# A Systematic Review and Meta-Analysis on Catastrophic Cost incurred by Tuberculosis Patients and their Households

**DOI:** 10.1101/2021.02.27.21252453

**Authors:** Ramy Mohamed Ghazy, Haider M. El Saeh, Shaimaa Abdulaziz, Esraa Abdellatif Hammouda, Amira Elzorkany, Heba Kheder, Nardine Zarif, Ehab Elrewany, Samar Abd ElHafeez

## Abstract

**Background:** As one of the World Health Organization (WHO) End Tuberculosis (TB) Strategy is to reduce the proportion of TB affected families that face catastrophic costs to 0% by 2020. This systematic review and meta-analysis aimed to estimate the pooled proportion of TB affected households who face catastrophic cost.

**Method:** A search of the online database through September 2020 was performed. A total of 5114 articles were found, of which 29 articles got included in quantitative synthesis. Catastrophic cost is defined if total cost related to TB exceeded 20% of annual pre-TB household income. R software was used to estimate the pooled proportion at 95% confidence intervals (CIs) using the fixed/random-effect models.

**Result:** The proportion of patients faced catastrophic cost was 43% (95% CI 34-52, I^2^ = 99%); 32% (95% CI 29 – 35, I^2^ = 70%) among drug sensitive, and 80% (95% CI 74-85, I^2^ = 54%) among drug resistant, and 81% (95%CI 78-84%, I^2^ = 0%) among HIV patients. Regarding active versus passive case finding the pooled proportion of catastrophic cost was 12% (95% CI 9-16, I^2^ = 95%) versus 42% (95% CI 35-50, I^2^ = 94%). The pooled proportion of direct cost to the total cost was 45% (95% CI 39-51, I^2^ = 91%). The pooled proportion of patients facing catastrophic health expenditure (CHE) at cut of point of 10% of their yearly income was 45% (95% CI 35-56, I^2^ = 93%) while at 40% of their capacity to pay was 63% (95% CI 40-80, I^2^ = 96%).

**Conclusion:** Despite the ongoing efforts, there is a significant proportion of patients facing catastrophic cost, which represent a main obstacle against TB control.

**PROSPERO registration:** CRD42020221283

## Introduction

Tuberculosis (TB) infection is one of the top 10 causes of death. It caused 1.2 million deaths in 2019. TB affects about one-quarter of the world’s population[1]. According to World Health Organization (WHO) report in 2020, WHO region that reported the highest incidence of TB was Africa region (266/10^5^) corresponding to 2.5 million cases. The South-East Asian region ranked the second (217/10^5^) corresponding to 4.3 million cases followed by the East Mediterranean region (114/10^5^) corresponding to 819 thousand case, and by Western Pacific region (93/10^5^) corresponding to 1.8 million cases. On country-based ranking, number of reported new cases is the highest in India (26%), Indonesia (8.5%), China (8.4%), Philippines (6.0%), Pakistan (5.7%), Nigeria (4.4%), Bangladesh and South Africa (3.6% for each). [2]

On 26 September 2018, WHO’s End TB Strategy was set and agreed by United Nation to end TB epidemic by 2030, with step wise milestones for 2020, 2025, and 2030. One of these Strategies is to reduce TB incidence rate and deaths by 90% and 95% respectively. It was also recommended to find TB missing cases by “active case finding (ACF) instead of passive case finding (PCF). ACF means systematic identification and screening of people with presumptive TB, in high-risk groups, using tests, examinations or other procedures that can be applied rapidly”, while PCF entails visiting health services for diagnosis[3, 4]. In addition, all TB patients or families should not suffer from catastrophic total costs (CTC) due to TB as one of the main obstacles for TB patients to complete their treatment; [5]. Catastrophic cost is defined as the total direct and indirect costs that reaches or exceed 20% of the pretreatment patient or household’s annual income. [5]of note, factors that aggravate this catastrophic cost are patient age and sex, socioeconomic status, Human immuno-deficiency virus (HIV) co-infection, and being infected with multidrug-resistant TB (MDR-TB) that does not respond to at least Isoniazid and Rifampicin, the 2 most powerful anti-TB drugs [6] [7].

The nominator of catastrophic cost is the summation of direct and indirect costs. The direct cost includes either medical cost (consultation fees, diagnostic tests and treatment) or non-medical cost (transportation, accommodation, increased food needs). Indirect cost includes lost wages due to unemployment; time spent away from work and associated loss of productivity. Moreover, patients also incur large costs in the pre-treatment phase to cover consultations and laboratory tests, symptomatic treatment, antibiotics trial, and hospitalization [8]. An important segment of the financial hardship is dissaving which means reduced financial strength of a household or engage the household in damaging financial coping strategies. This will reduce the financial capacity and their coping with the financial shocks and cast them into the poverty trap. [9] Dissaving can take many forms like taking out a loan, taking children out of education, selling assets, reducing consumption to below basic needs to cope with health-related expenditure [8–10].

Consequently, WHO developed the TB patient cost survey to properly assess the total costs and proportion of patients facing catastrophic cost. This tool provide a standardized methodology for cross-sectional surveys in TB affected countries [11]. Many studies used this cost survey to report catastrophic cost, catastrophic health expenditure, or hardship financing incurred by TB patients [12–14]. Some literatures calculated catastrophic cost for drug sensitive, MDR or HIV co-infection [14–16]. Other studies estimated compared this cost considering adoption of different case finding strategies (ACF versus PCF) [17, 18]. In response to this reported catastrophic cost, the Global TB Program endorses social protection initiatives to complement Universal health coverage (UHC) initiatives [19, 20]. Examples of social protection interventions include cash transfers, food assistance, disability grants and health insurance. Those global financial supports already exist in most countries, but may not be fully implemented [7].

At the end, keeping in mind that COVID-19 pandemic may reverse the achieved progress in the TB control as many countries directed their resources toward pandemic containment. In addition, there are no published systematic reviews that report the pooled proportion of patients suffering from catastrophic cost; we aimed to perform this systematic review and meta-analysis to estimate the proportion of catastrophic cost among TB patients and their households in attempt to support the ongoing TB control programs.

## Method

This systematic review and meta-analysis was conducted according to the Preferred Reporting Items of the Systematic Reviews and Meta-Analyses (PRISMA) guidelines [21].

### Data source and search strategy

EMBASE, Scops, EBSCO, MEDLINE central/PubMed, ProQuest, Scielo, SAGE, Web of science, and Google scholar databases were searched for articles without timeframe, geographical or language restrictions up to November 20^th^, 2020 by two authors (ShA & NZ) then revised by (RMG& SA). Highly focused and sensitive search strategies were developed by RMG after the approval of PubMed Help Disk. The search terms include (“tuberculosis” OR “Mycobacterium tuberculosis” OR “Koch’s disease” AND “catastrophic cost”). References from relevant studies were screened for supplementary articles.

### Study selection and data extraction

We aimed to include observational studies, which reported the proportion of patients suffering from catastrophic cost during the intensive (first 2 or 8 months of treatment in DS or MDR respectively) or the continuation phases of TB treatment.

The primary endpoint of interest was the proportion of TB affected patients and their households who face catastrophic cost. It was defined as the total direct and indirect costs due to TB reaches or exceed 20% of the patient or household’s annual income [5]. Furthermore, CTC was assessed among patients according to their drug sensitivity as DS or MDR (with or without HIV), and strategy of case finding (ACF versus PCF).

Secondary outcomes were the proportion of the direct to the total cost of TB among DS or MDR, with or without HIV, catastrophic health expenditure CHE (defined as direct cost that reaches or exceeds 40% of patients capacity to pay or 10% of their household income [22], and the different coping strategies.

Titles and abstracts were screened independently by four authors (AM, ShA, NZ, and EE), who discarded articles not pertinent to the topic. Non-observational studies, case reports, editorial, reviews, letters, and studies that estimated the direct and indirect cost of the population as a one unit not individually were excluded from qualitative analyses but screened for potential additional references. Three other authors (RMG, SA & HE) solved the discrepancies on study judgements. Data extraction and analysis were performed by (RMG, AM, HE) and independently verified by (SA)

### Data analysis

The proportion of CTC among TB patients was pooled using the random-effects model. To ensure robustness of the model and susceptibility to outliers, pooled data was also analyzed with the fixed-effects model. Heterogeneity was assessed by the Chi-squared test on N-1 degrees of freedom, with an alpha of 0.05 considered for statistical significance and the Cochrane-I-squared (I^2^) statistic. I2 values of 25%, 50% and 75% were considered to correspond to low, medium and high levels of heterogeneity, respectively.

Sources of heterogeneity, for identifying possible effect modifiers on the pooled analyses, were explored using:

1. Sensitivity analysis (leave one out sensitivity analysis, GOSH sensitivity analysis, remove outliers)
2. Subgroup analysis: we categorized the catastrophic cost at 20% for ACF and PCF patients according to country where studies were conducted (inside/outside) India.
3. Met-regression: The impact of country where the survey was conducted (high versus low incidence of TB) [23], quality of the study, sex, and population criteria (drug sensitivity, drug resistant with or without HIV) on the size effect of studies to explain the substantial heterogeneity. The forest plot was used to visualize the degree of variation between studies. All data analysis was performed R software version 4.0.3 using Harrer hand-on guide [24].

### Publication bias

Publication bias was investigated by visual inspection of funnel plots, and by Egger’s regression test.

### Quality assessment

The Newcastle-Ottawa Scale (NOS) was used to assess the quality of studies. Studies were classified according to the NOS as: very good studies (9-10 points), good studies (7-8 points), satisfactory studies (5-6 points), and unsatisfactory studies (0-4 points).[25]

## Results

### Search results

The flow diagram of the selection process is shown in figure 1. In total of 5114 potentially relevant articles were found after data base search. One additional citation was found through a personal search, of this number, 1922 articles were excluded as duplicates by Endnote X8. After title and abstract screening 3041 article were excluded (201 duplicates found manually, 2840 irrelevant). Two unpublished data were included to the 152 text eligible articles to full text screening, in addition we added 2 articles were added manually. A total of 29 articles were therefore reviewed in detail and included in the analysis. The main characteristics of these studies are summarized in table 1. The inter-rater agreement for inclusion was κ=0.95 and for the quality assessment was κ=0.84

**Figure 1:**
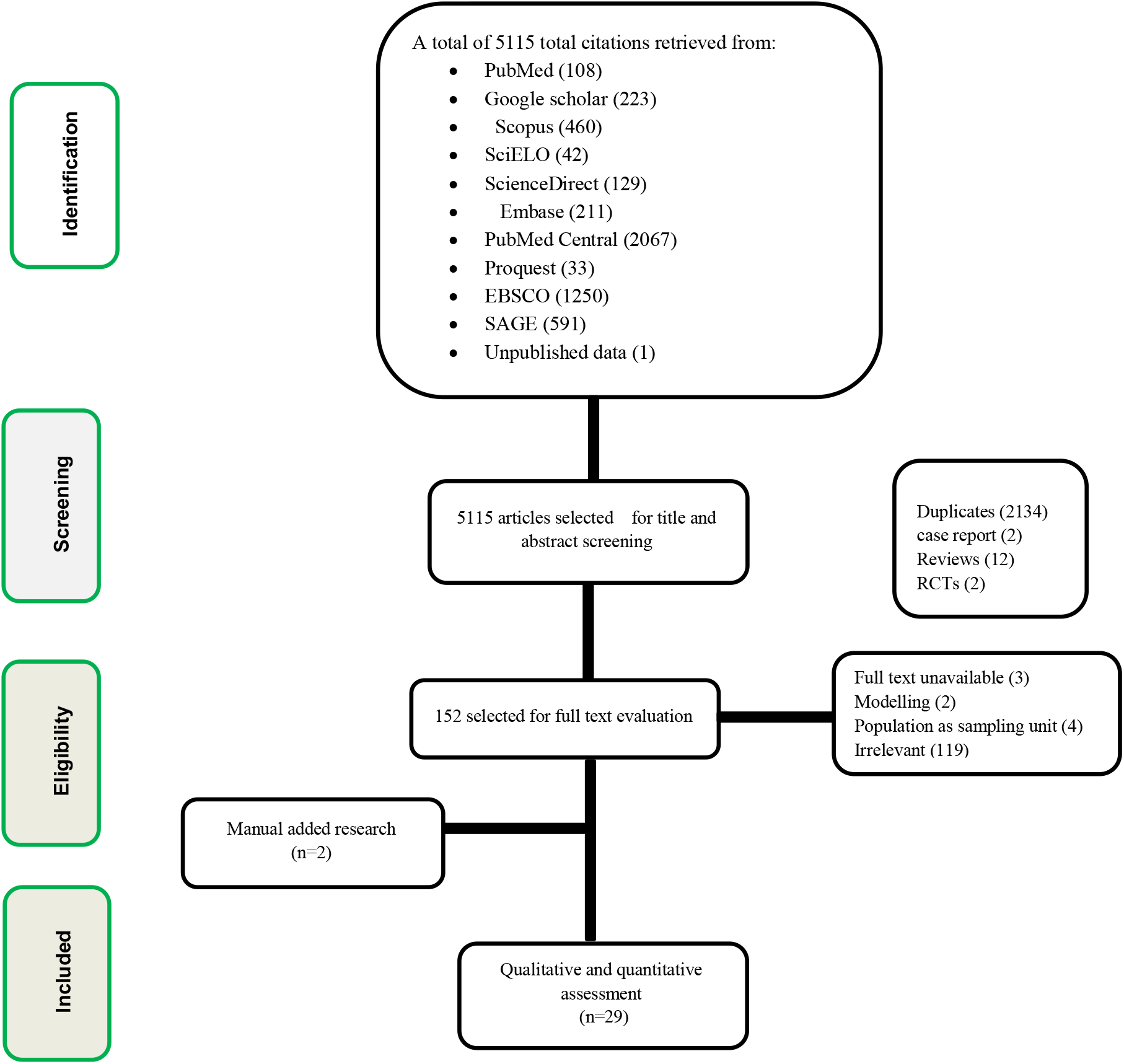
PRISMA flow charts of studies included in meta-analysis of catastrophic cost/expenditure among patients with tuberculosis.

### Study characteristics

Qualitative synthesis included 29 studies conducted in 15 countries; six studies from India, five from China, four from Indonesia, one study from each of the following countries (Egypt, Zimbabwe, Nepal, Lao PDR, Ghana, Pakistan, Vietnam, Cambodia, Peru, and Cavite), and two studies from each Uganda, and South Africa. Of included studies there were 5 cohort studies [12], [13], [17], [26] &[27]. One mixed methods study [28], while the other 23 studies were cross-sectional. Male sex presentation ranged from 30% [29], to 77% [18]. The sample size ranged from 50 [29], to 1178 [30]. The tool used for estimation of the cost survey were either WHO TB cost survey tool, [5], [15], [30], [31], [32], [33], [34], [35], [36], [37], [38], [39], [40] & [44], or adapted WHO tool to Indonesian context [12] & [41], or structured questionnaire [17], [26], [42], [43], pre-coded interview scheduled [18],or tool of stop TB partnership, [13], [27], [28],or headcount tool [44], or Lumley T. survey [14], or TB coalition tool [16]. On the other hand, there were two studies not mentioned the tool used [29], [45]. The percent of patients facing catastrophic cost at cut off point 20% ranged from 4% in study of Mihir et al, study [46], to 87% in study of Wang et al, [44]. Regarding the percent of MDR-TB patients that facing catastrophic cost, they ranged from (68%), reported by Mullerpattan 2019 to (90%), reported by Collin et al, 2018 however DS-TB patients ranged from 24%, in the study of Gadallah,2018 to 42%, in the study of Rebecca L.Walctt, 2020. The percent of ACF patient facing catastrophic cost ranged from 9% to 44%, however the percent of PCF patient ranged from 29% to 61% [18] & [39]. Hardship financing was discussed only in two studies[14] [45]. Seven studies discussed coping cost [16], [27], [30], [33], [35], [37] & [45]. Regarding the quality score, it was ranged from (3-unsatisfactory) [29] to (9-Very good) [34]. The Good score ranged from 7 to 8 pints, was among thirteen studies [5], [16], [17], [26], [28], [31], [32], [33], [37], [38], [39], [41] & [45]. While Satisfactory score which ranged from 5 to 6 points, was illustrated in the remaining fourteen studies, [12], [13], [14], [15], [18], [27], [30], [35], [36], [40], [42], [43], [44] & [45].

### Publication bias

The 29 studies reported the catastrophic cost at 20% were be assessed for the risk of bias by the funnel plot and Eggers’ test [t = −1.188, P-value= 0.24], which revealed the absence of asymmetry and decline the presence of publication bias. **Figure 2**

**Figure 2:**
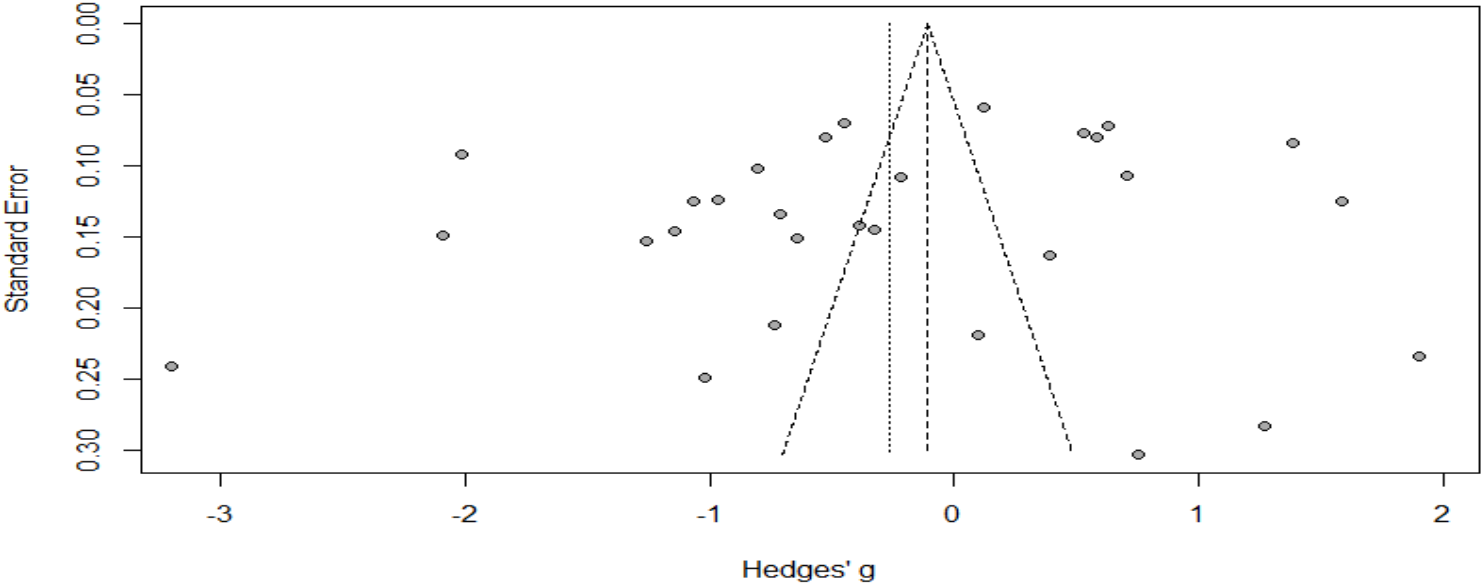
Funnel plot of studies included in estimation of the proportion tuberculosis patients facing catastrophic cost.

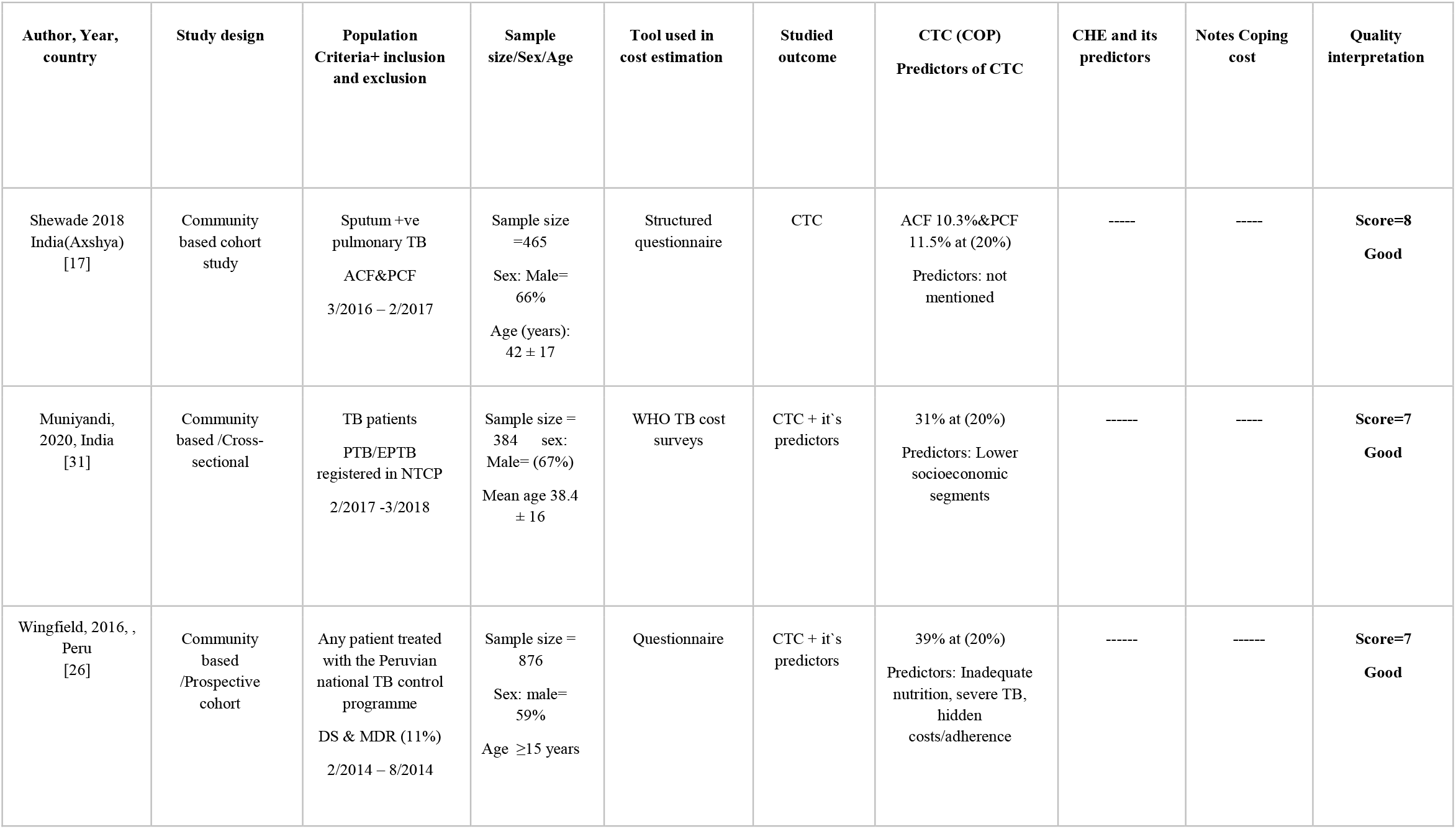
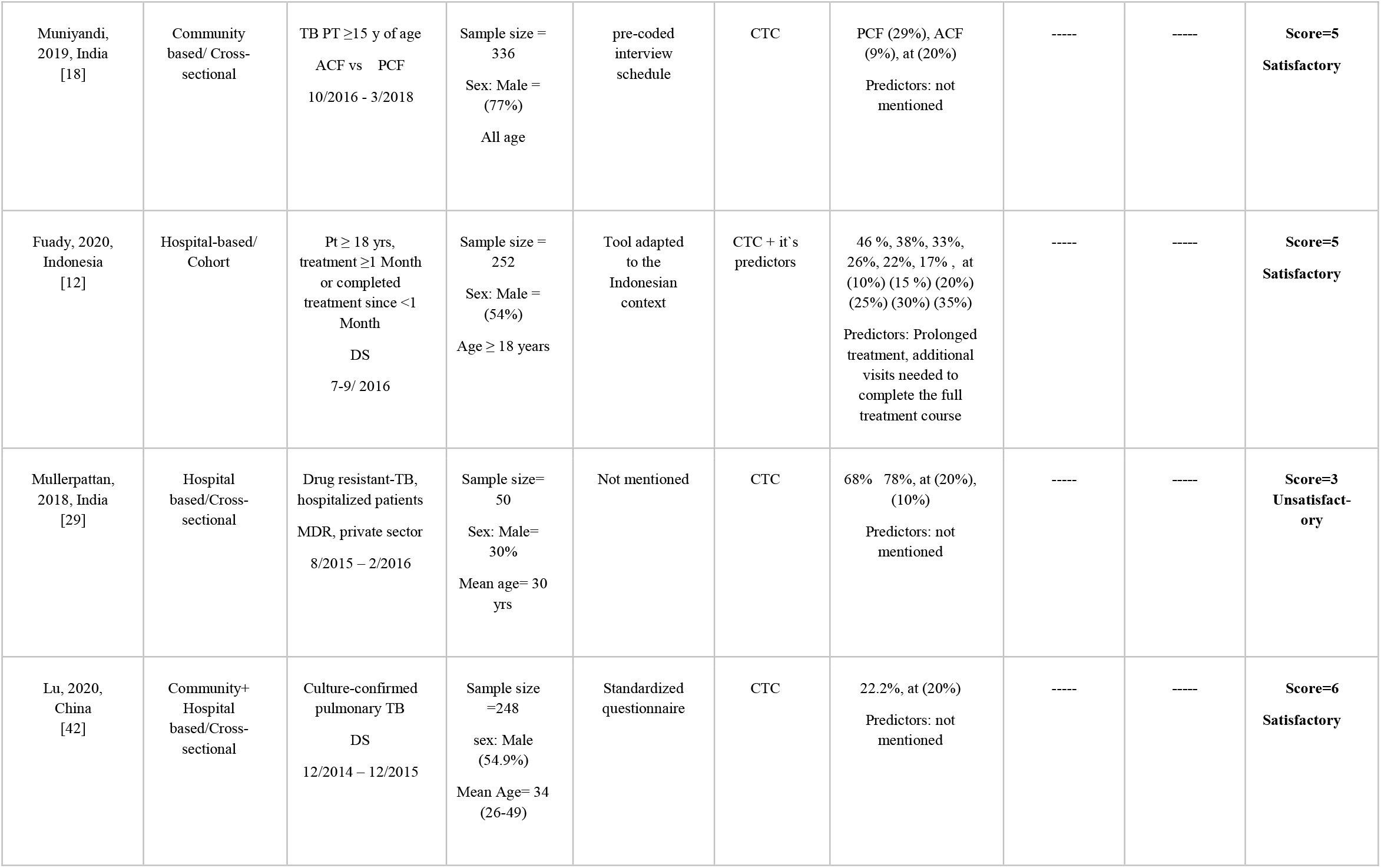
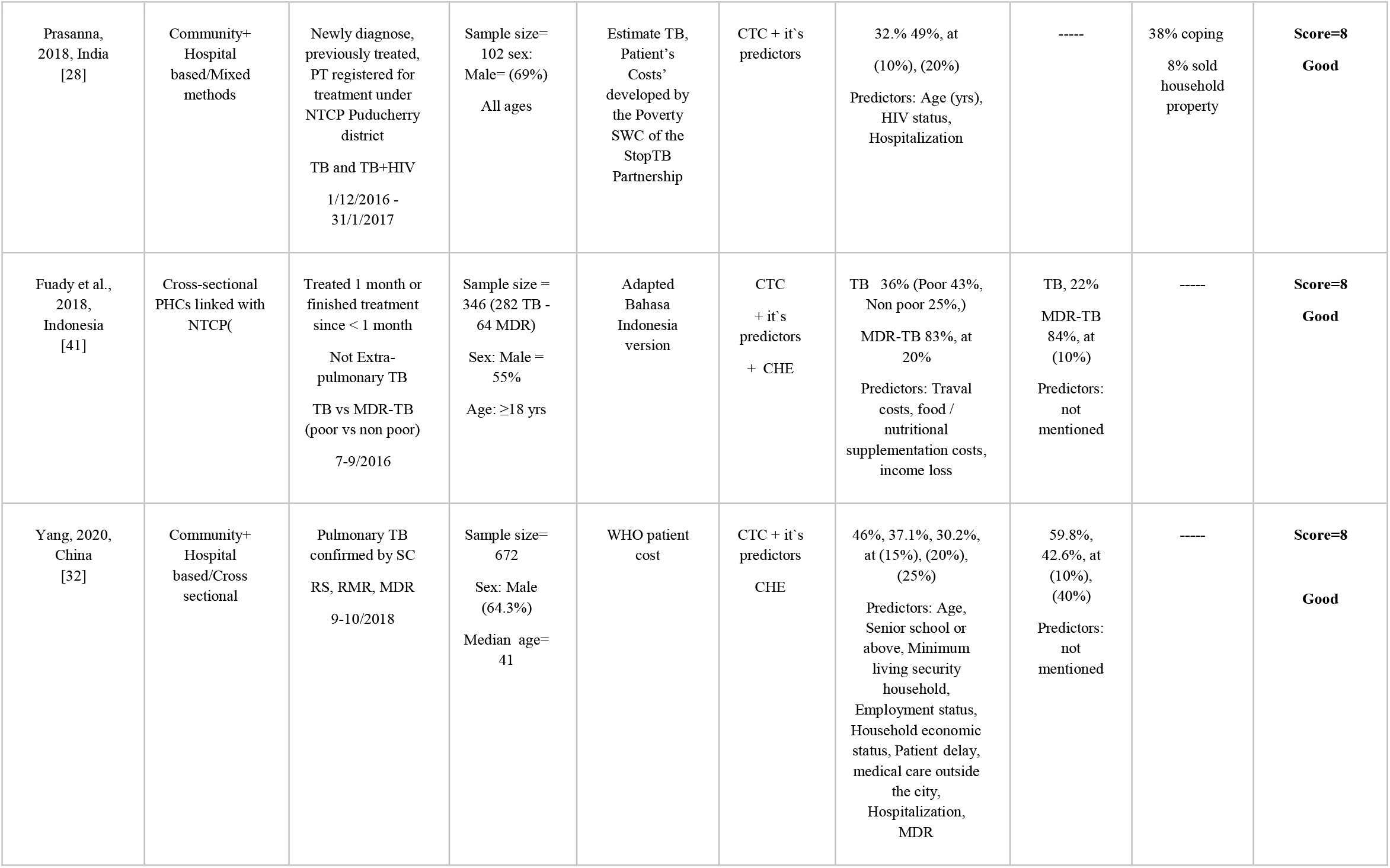
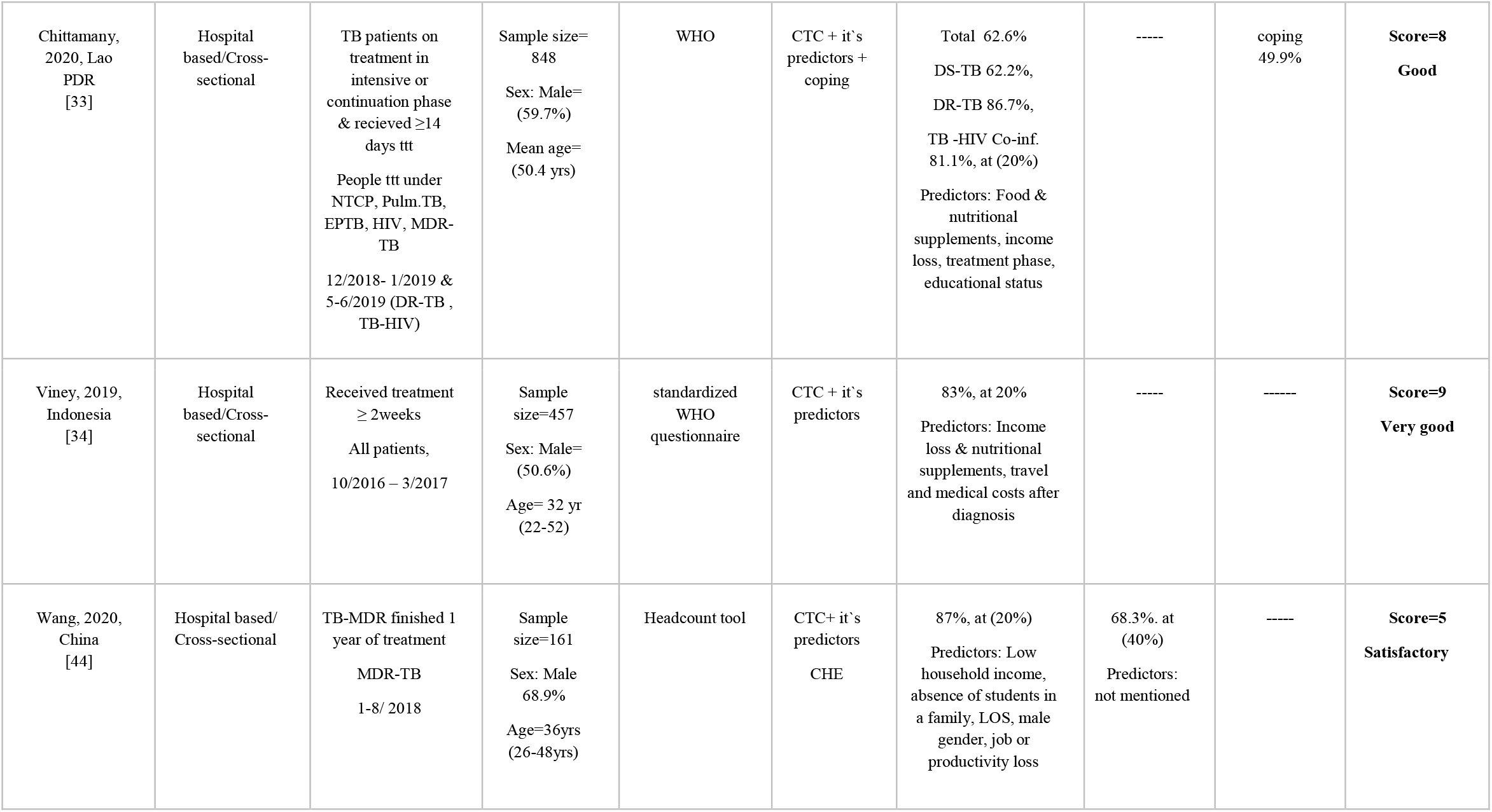
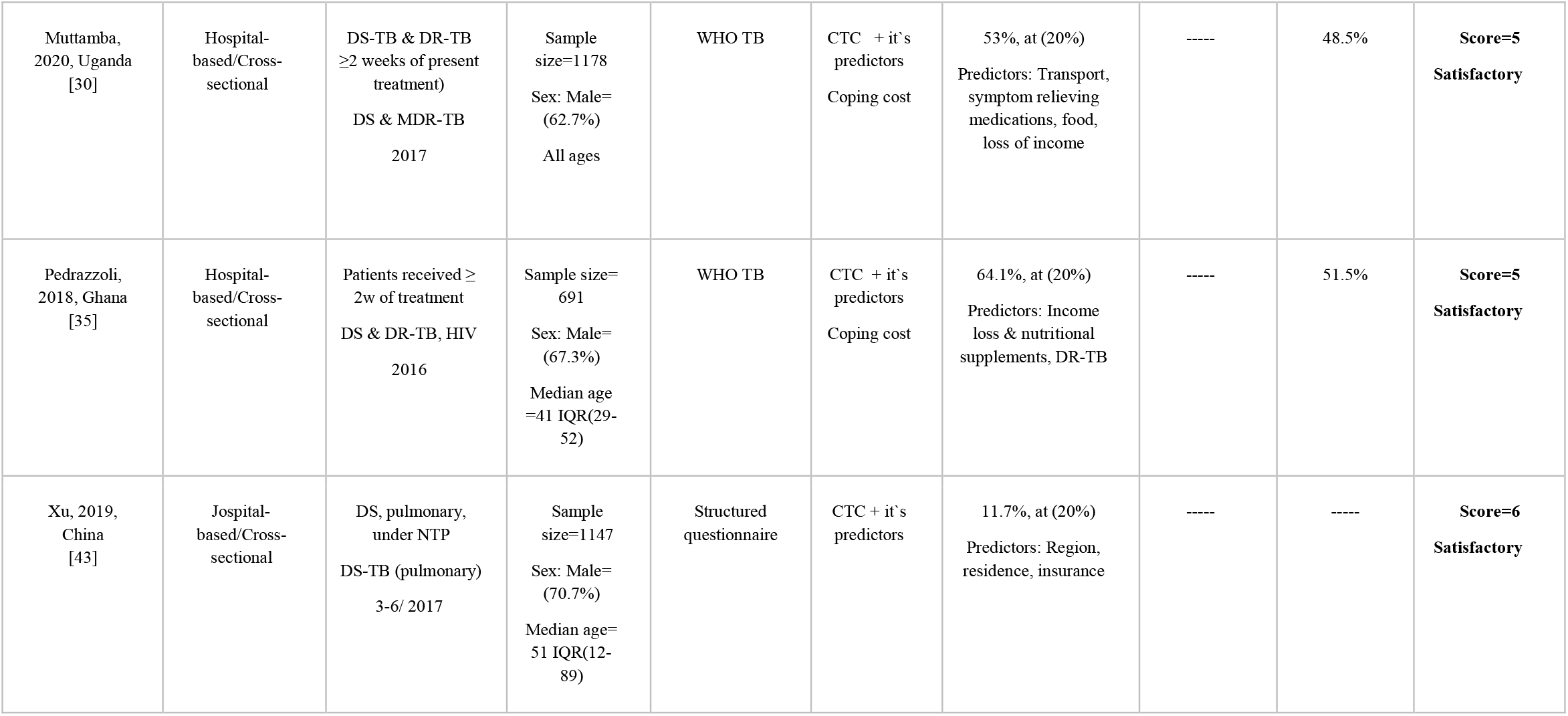
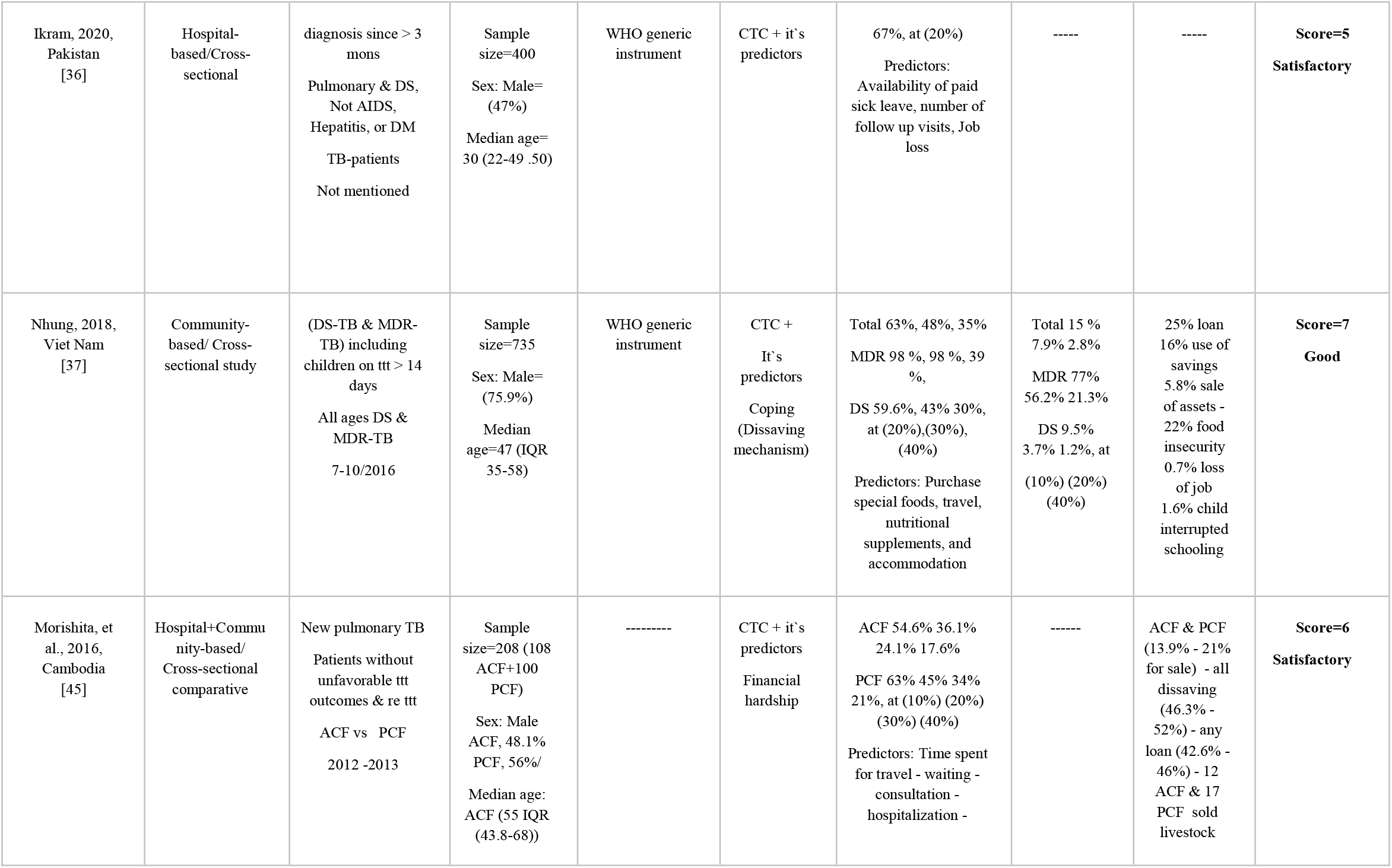
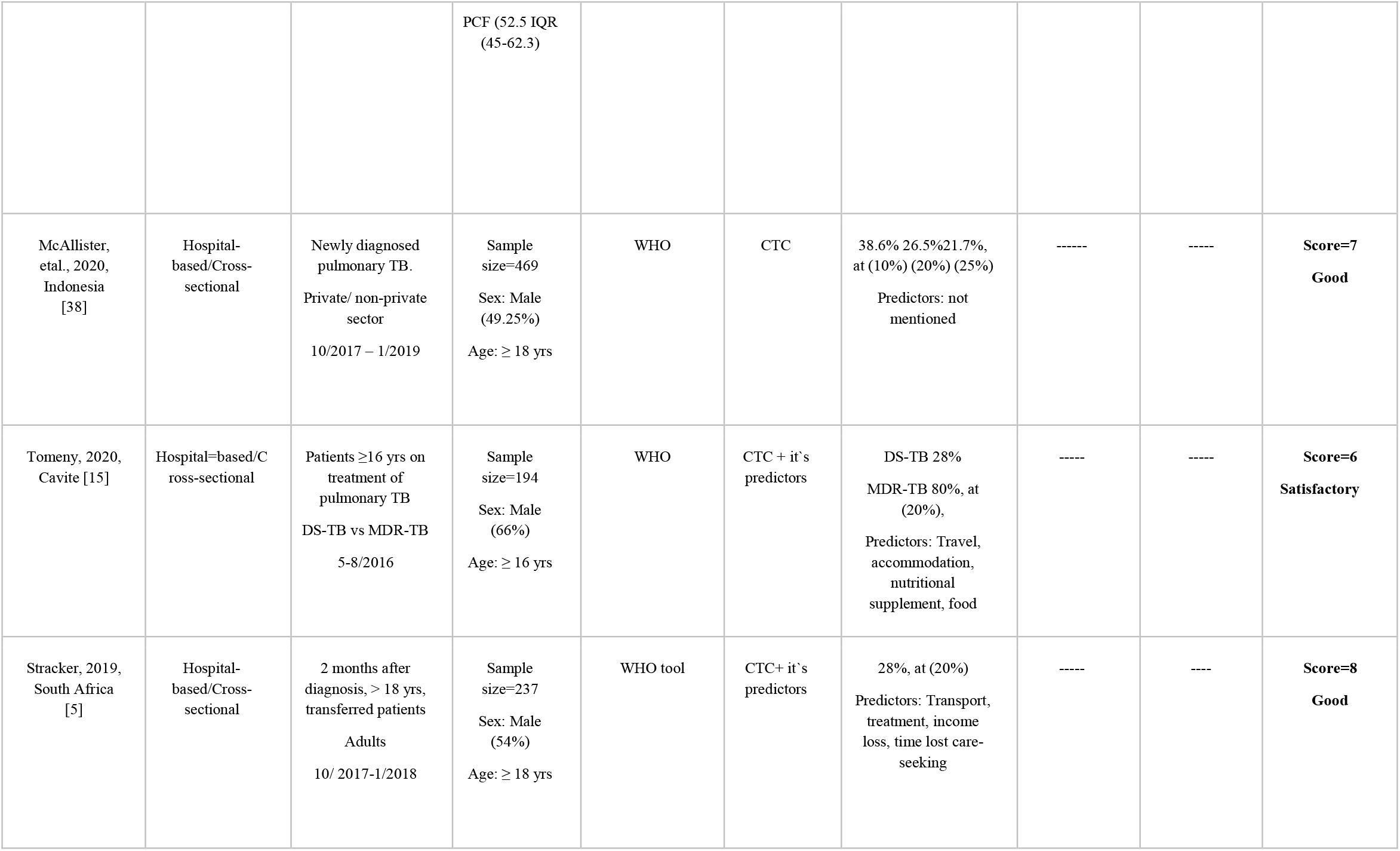
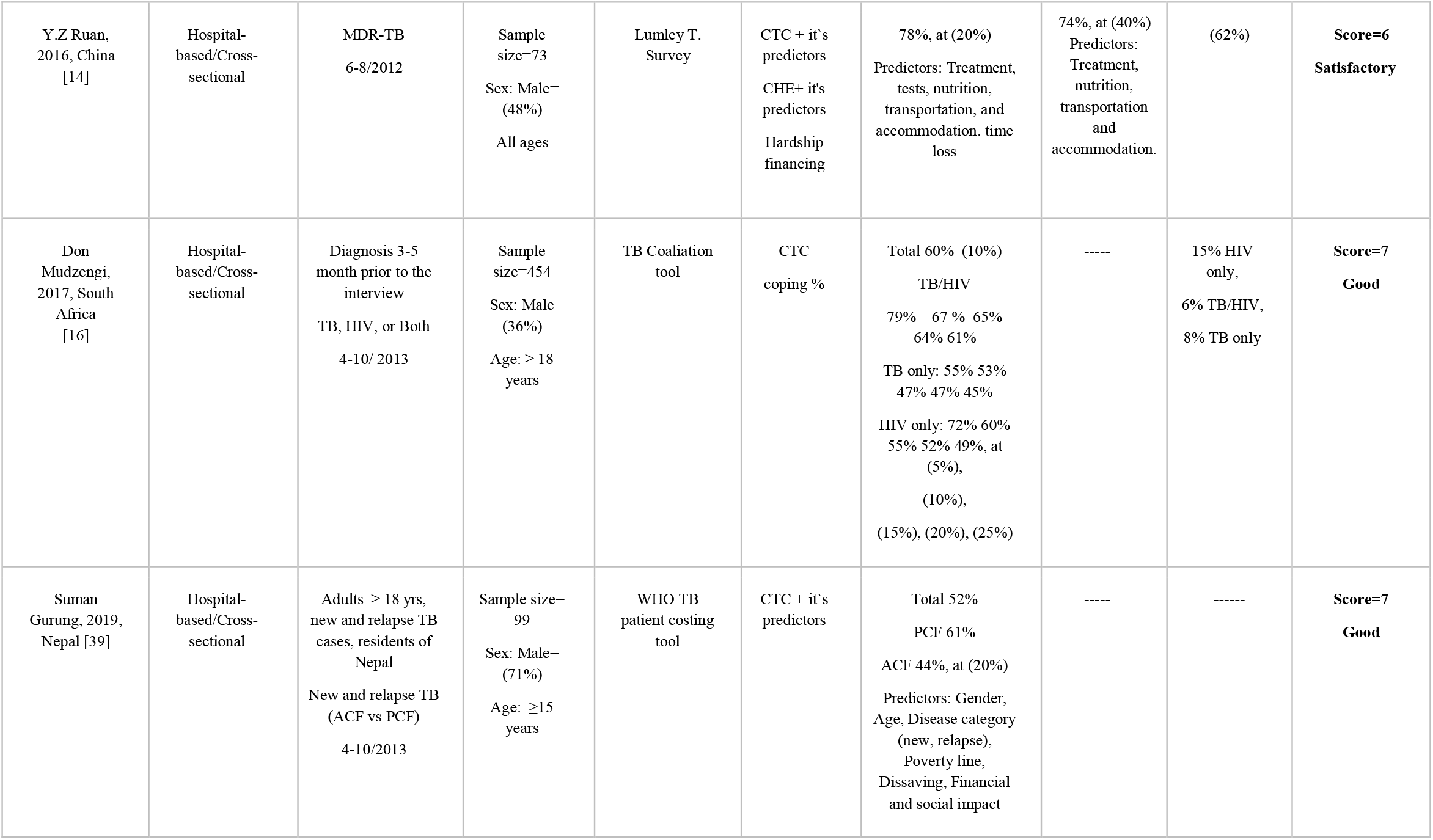
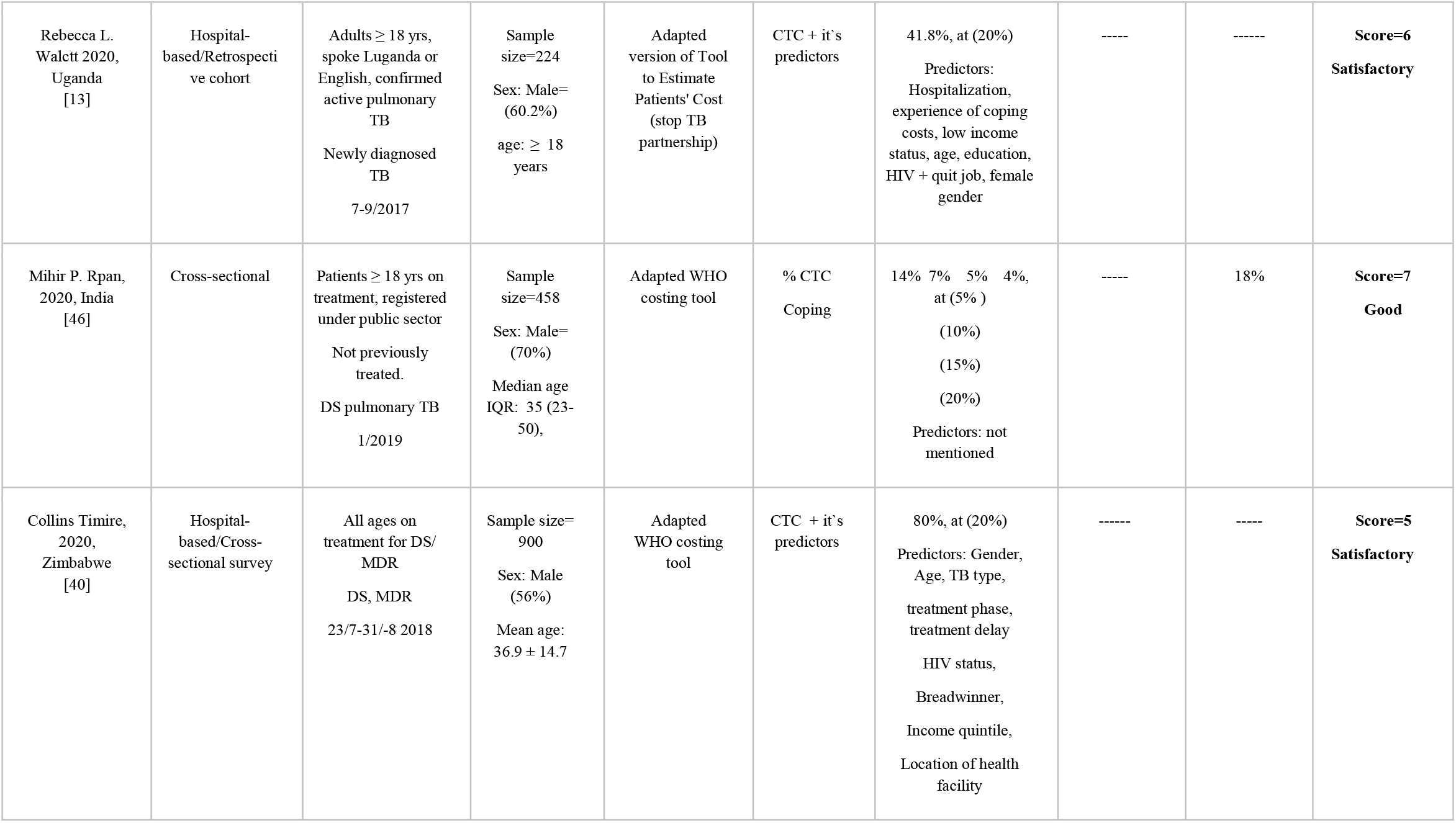
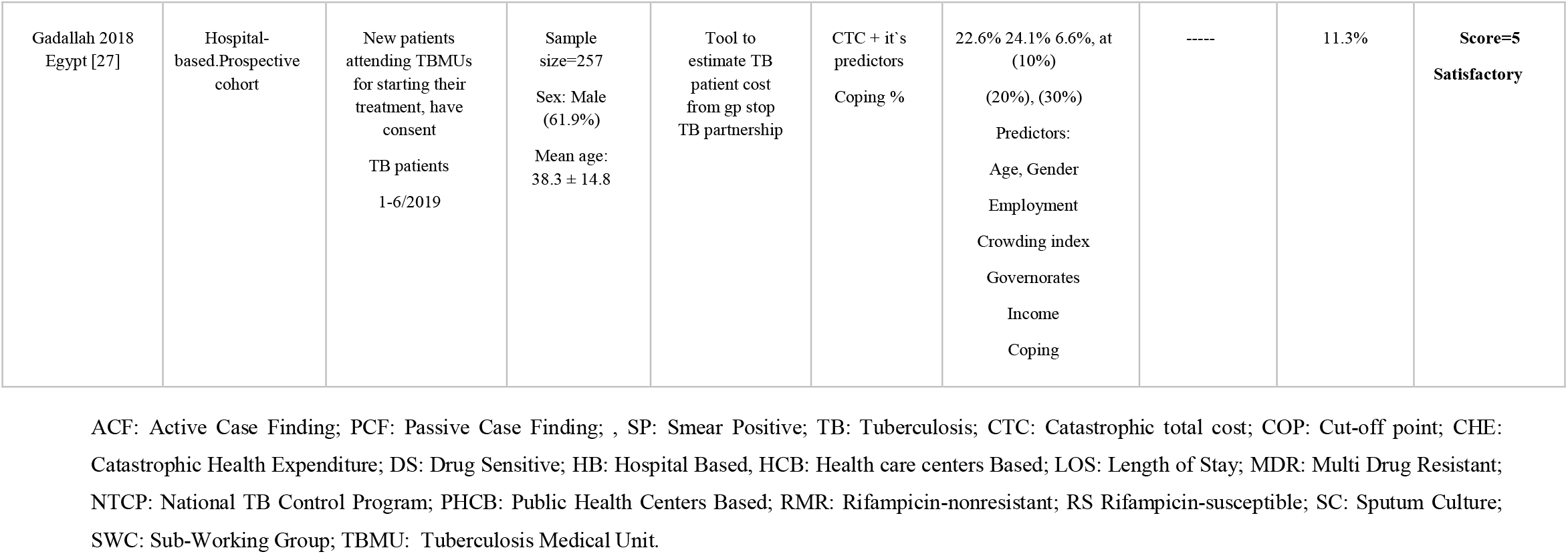

## 1. Primary outcome

### 1.1 Catastrophic cost at cut-off point 20%

The pooled prevalence of catastrophic cost among 11750 TB patients included in 29 studies at cut-off point of 20% was 43% (95% CI:34-52) with high heterogeneity (I^2^ = 99%). **Figure (3)** To identify the cause of this substantial heterogeneity we conducted meta-regression. Predictors were sex, country where the study conducted (had high incidence vs none)[23], drug sensitivity (DS or MDR± HIV), and quality of the study. The model was significant P<0.0127, R^2^=51.57%. This model explained more than 50% of the reported heterogeneity. The identified predictors country (high vs low incidence) (β=−0.194, P=0.04) and type of patients regarding drug sensitivity (DS or MDR) and HIV co-infection (β=0.289, P=0.026).

In this study, there are multiple main predictors of catastrophic cost like food and nutritional supplements [33–35],travel and transportation [30, 32, 45], age category [26, 28, 32], employment status [26, 32, 36, 41, 44], the socioeconomic status[13, 26, 27, 32, 41, 44, 47], MDR or HIV positive [28, 32, 35, 47], male gender, [26, 27, 44], and duration of hospitalization [13, 28, 32, 44, 45].

**Figure (3).**
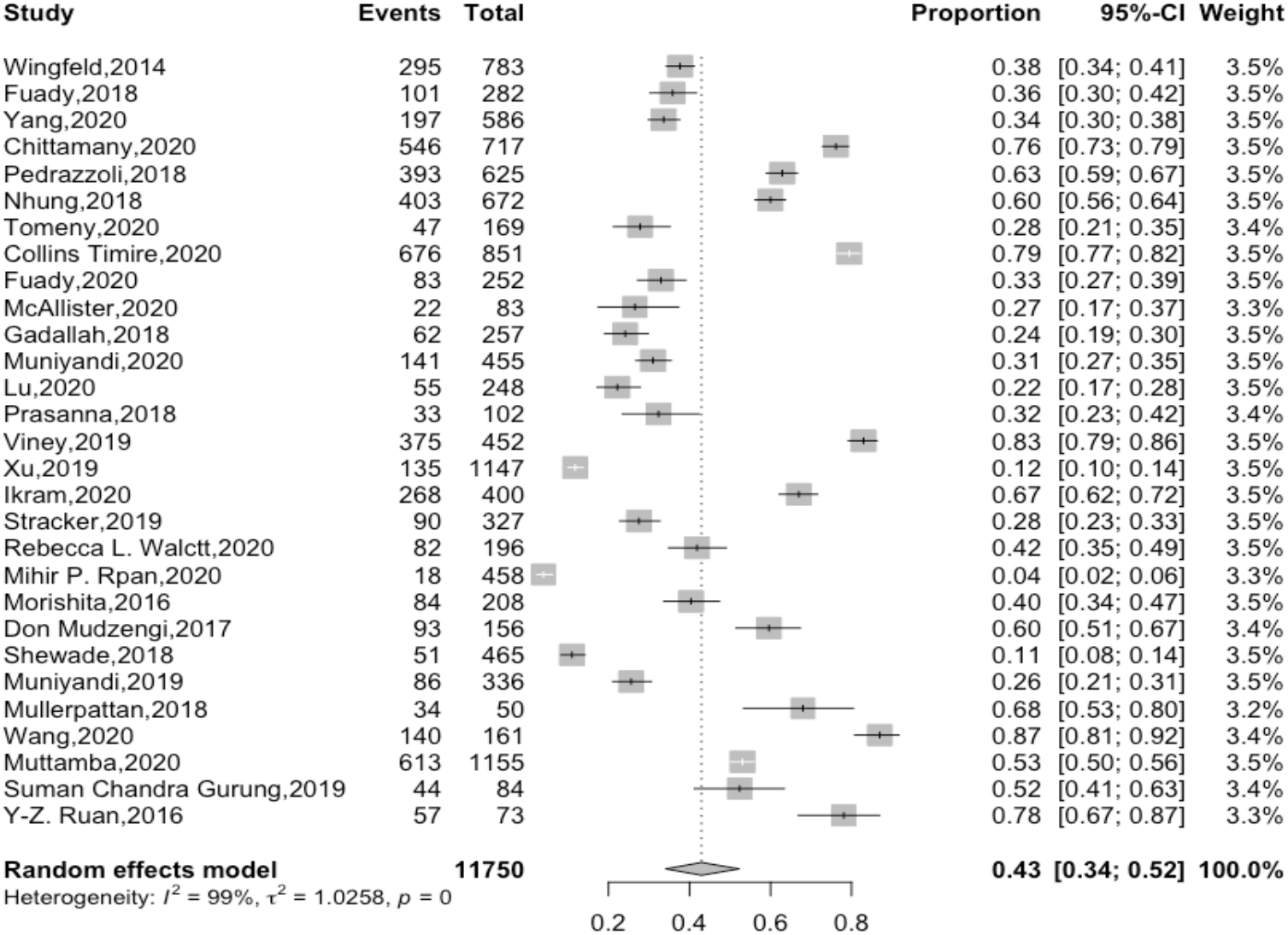
Pooled proportion of catastrophic cost at 20%.

### 1.2 Coping strategy

In response to balance the enormous financial burden they encounter, the TB-affected families may adopt some coping strategies. Borrowing money, taking out loans, pledging gold and jewels, bringing their children out of schools or selling assets are options to compensate the income loss and the high out-of-pocket expenses [37, 45]. All these approaches are referred to as “dissaving” which is the core of the hardship financing dilemma.

### 1.3 Pooled proportion of catastrophic cost at 20% among different subgroups

#### 1.3.1 Pooled proportion of catastrophic cost at 20% among TB drug sensitive

The pooled proportion of patients facing catastrophic cost was 39%, 95CI (28-51%), the reported heterogeneity was 99%. After removing outliers, the pooled proportion of 11 studies recruited 3492 patients dropped to 32%, 95% CI [29 – 35]. The pooled prevalence of DS-TB patients facing catastrophic costs ranged from 24%, 95%CI [19 – 30] in the study of Gadallah, 2018 [27] to 42%, 95% CI [35 – 49] in the study of Rebecca L.Walctt, 2020 [13].The heterogeneity of the included studies was as follows; I^2^ = 70%, P < 0.01. **Figure (4)**

**Figure (4).**
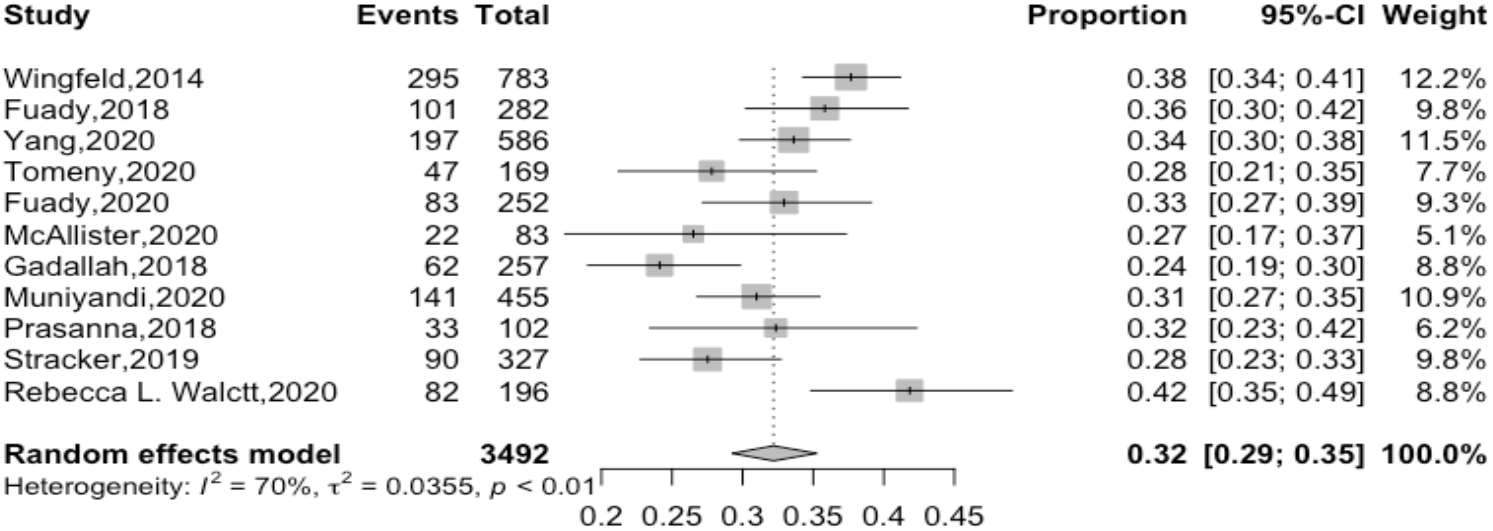
Pooled proportion of catastrophic cost at 20% among drug sensitive after removing outliers.

#### 1.3.2 Pooled prevalence according to TB drug resistant

With a heterogeneity of 92%, the pooled proportion of TB affected household of MDR patients facing catastrophic cost among 1879 patients was 78%, 95%CI, [86%-86%]. After removing outliers, the pooled proportion of patients facing catastrophic cost among 574 patients with MDR reached 80% 95%CI [74-85%], I^2^=54%. The highest proportion (90%) reported by Collin et al, 2018[40], while the lowest proportion (68%) reported by Mullerpattan 2019 [29]. **Figure (5)**

**Figure (5).**
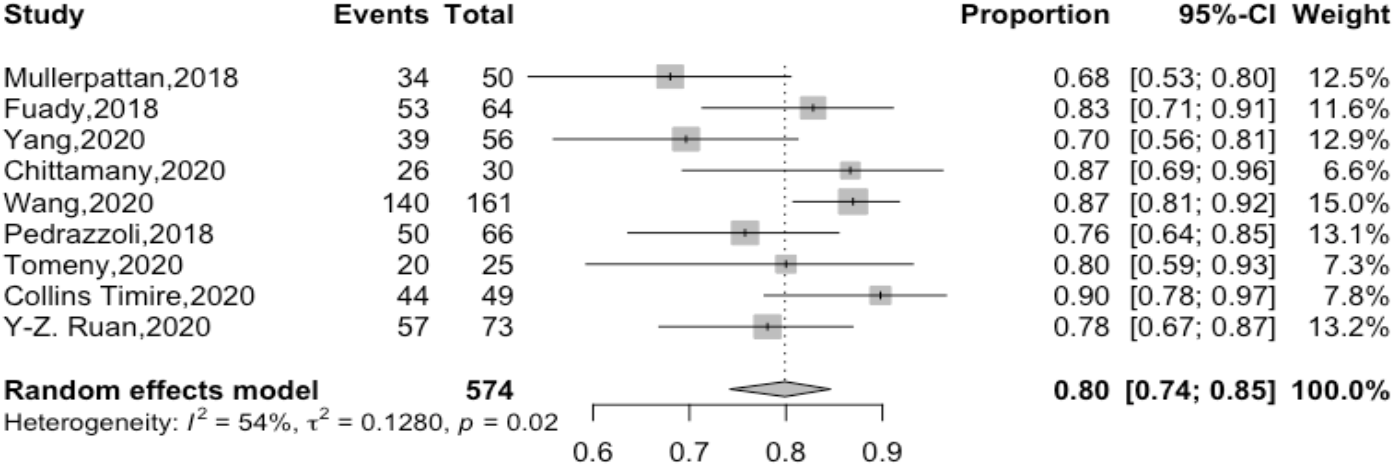
Pooled proportion of catastrophic cost at 20% among drug resistant after removing outliers.

#### 1.3.3 Pooled proportion of TB-HIV co-infected patients facing catastrophic cost at 20%

The pooled proportion of 796 TB patients with HIV facing catastrophic cost at 20% was 76%, 95%CI [65-85%], with a heterogeneity of 88%. After conducting leave-one out sensitivity analysis, the study of Don Mudzengi et al 2017 [16], removed. The heterogeneity dropped to 0% and the pooled proportion patients facing catastrophic cost has increased to 81%, 95%CI [78 – 84] as it illustrated in **Figure (6)**

**Figure (6).**
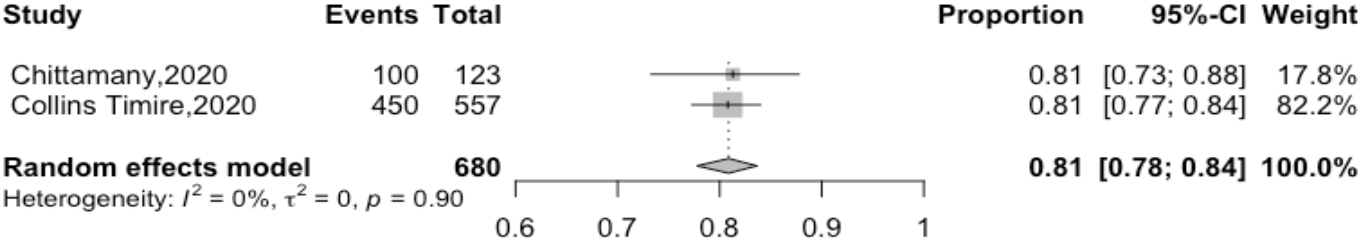
Pooled proportion of catastrophic cost at 20% among TB and HIV infected patients after removing outlier.

**Figure (7).**
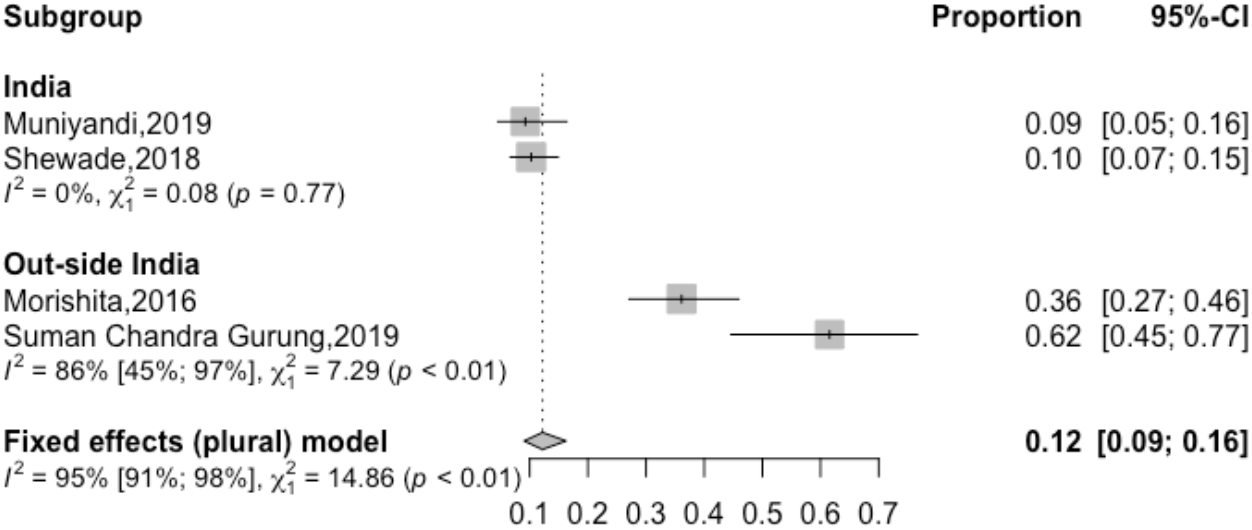
Pooled proportion of catastrophic cost at 20% among during active case finding after sub-group analysis.

#### 1.3.4 Pooled proportion of TB facing catastrophic cost at 20% through active case finding (ACF)

The proportion of patients facing catastrophic cost among 491 patients exposed active case finding ranged from 9%, 95%CI [7-15%] to 62%, 95%CI [45-77%]. After subgroup analysis based on the country where the ACF was implemented (inside/outside India). The pooled proportion was 10% 95%CI [7-14%], I^2^= 0% inside India and 48%, 95CI(25-72%) I^2^ 86% outside India.

#### 1.3.5 Pooled proportion of TB facing catastrophic cost through passive case finding (PCF)

The proportion of patients facing catastrophic cost among 638 patients during passive case finding ranged from 12%, 95%CI [8-17%] to 45%, 95%CI [35-55%]. The pooled proportion was 42%, 95%CI [35-50%]; It is worthy to note that heterogeneity was 94%. We further subdivided the studies according to the studied country (inside/outside) India. The pooled proportion of TB household facing catastrophic cost was 19% 95CI (7-41%), I2=95% while outside India 45 95CI(37-53%), I2=0%. **Figure (8)**

**Figure (8).**
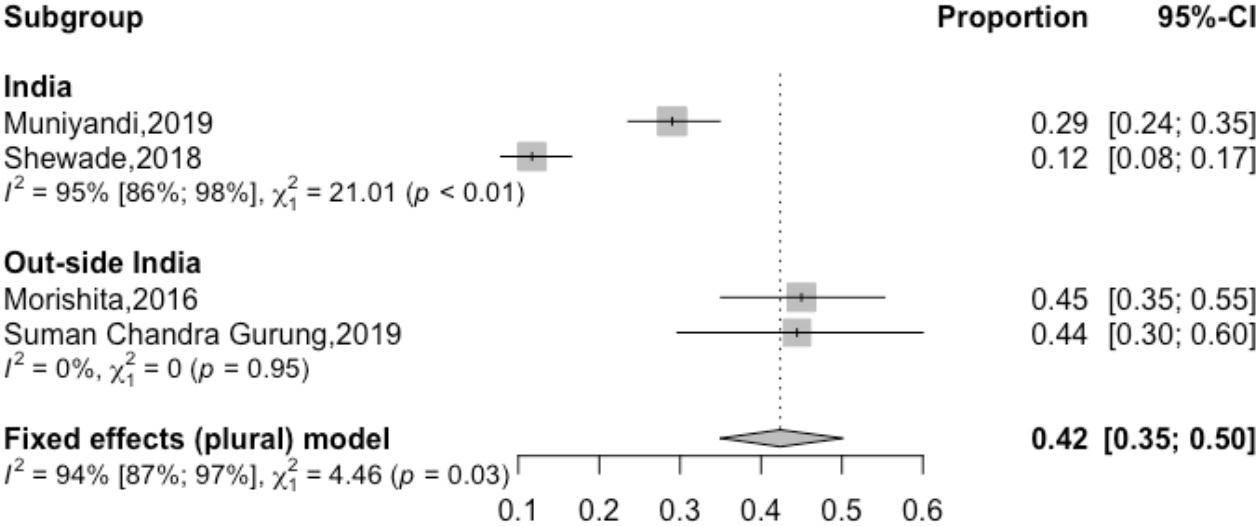
Pooled proportion of catastrophic cost at 20% among during passive case finding after sub-group analysis.

## 2. Secondary Outcome

### 1.1. Proportion of direct cost to the total cost

#### 1.1.1. Pooled prevalence according to drug sensitive

The proportion of the direct cost to the total cost addressed in 6 studies, the pooled proportion of direct to total cost at catastrophic cost of 20% was not calculated as the heterogeneity was high. The proportion was variants, two studies reported similar proportions, Tomeny, 2020[15] & Collins Timire, 2020 [47] with a proportion of 41% and 43% respectively. However, higher proportion 52% reported among Chittamany2020[33] and Nhung, 2018[37]. Two other extreme values reported, 33% by Fuady 2018 [41] and 65% reported by Muttamba, 2020 [30].

#### 1.1.2. Pooled proportion of direct cost in MDR

The proportion of the direct cost to the total cost at 20% addressed in 7 studies, ranged from 26% in Chittamany, 2020 [33] to 93% in Yang, 2020[32]. Low proportions were observed in Fuady, 2018[41], Tomeny, 2020 [15], and Collins Timire, 2020 [47] with proportion of 32%, 34% and 49% respectively, while high proportion also reported in Muttamba, 2020[30], with 66% and in Nhung, 2018[37] with 68%. The pooled proportion of direct to total cost was difficult to assess because of the heterogeneity which wasn’t explained even after a meta-regression performed.

#### 1.1.3. Pooled proportion of direct cost to total cost in case of active case finding (ACF)

The pooled proportion of the direct cost to the total cost was addressed in 3 studies, the pooled proportion of direct to total cost was 25%, 95%CI [16-37%], I^2^=83%. After conducting leave one out sensitivity analysis, the Gurung, 2019 [39], was removed, the pooled proportion dropped to 29%, 95%C1 [20-41%] I^2^=55%. **Figure (9)**

**Figure (9).**
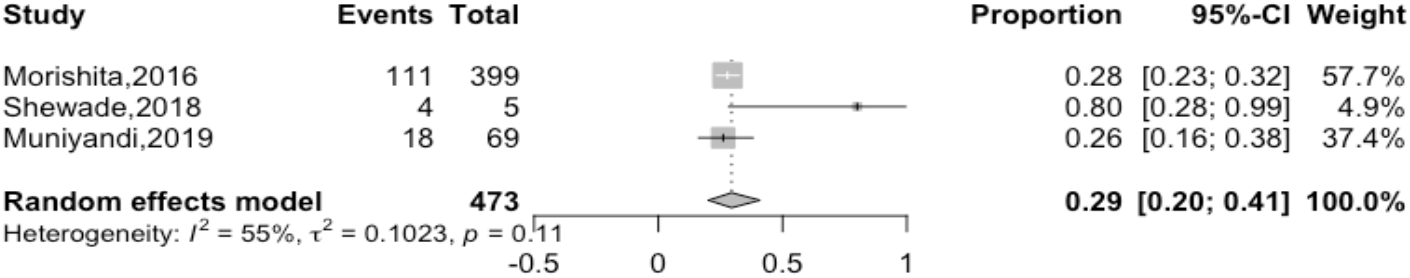
Pooled proportion of direct to total cost at catastrophic cost of 20% among active case finding after removing the outlier.

#### 1.1.4. Pooled proportion of direct cost to total cost in case of passive case finding (PCF)

The pooled proportion of the direct cost to the total cost addressed in 4 studies [17, 18, 39, 45], the pooled proportion of direct to total cost was 38%, 95%CI [32-46%], I^2^=83%. After conducting leave one out sensitivity analysis, the Shewade et al, 2018[17], removed, the pooled proportion dropped to 37%, 95%C1 [34-40%] I^2^=0%. **Figure (10)**

**Figure (10).**
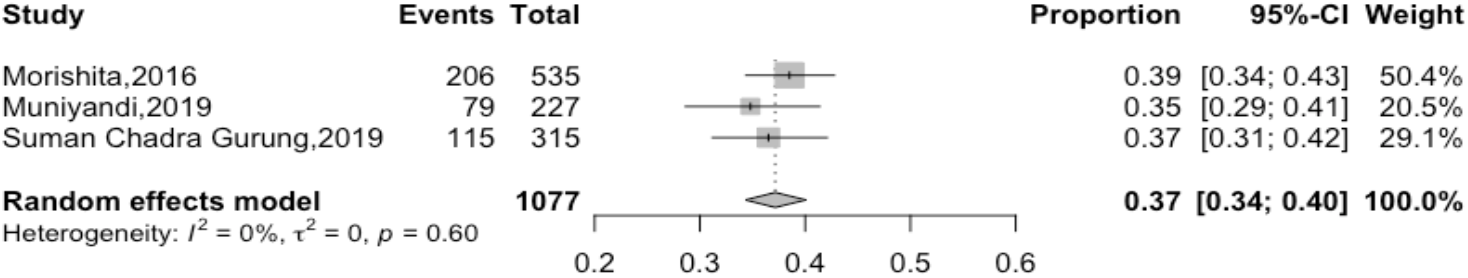
Pooled proportion of direct to total cost at catastrophic cost of 20% among passive case finding.

#### 1.1.5. Proportion of direct cost to total cost in case HIV and TB co-infection

The proportion of the direct cost to the total cost addressed in 2 studies. Don Mudzengi, 2017[16] and his team showed that the proportion of direct cost to the total cost was 30% among HIV and TB co-infection patients, while a higher proportion reported in Chittamany, 2020[33] with 59%. As we couldn’t pool the study because of the high un-explained heterogeneity.

The pooled proportion of the direct cost to the total cost addressed in 14 studies, the pooled proportion of direct to total cost was 55%, 95%CI [43-66%], I^2^ = 99%. After conducting outliers removal, the studies [16, 43, 44, 48–50] were excluded, the pooled proportion dropped to 51%,95%CI [43-59%], I^2^= 96%. **Figure (11)**

**Figure (11).**
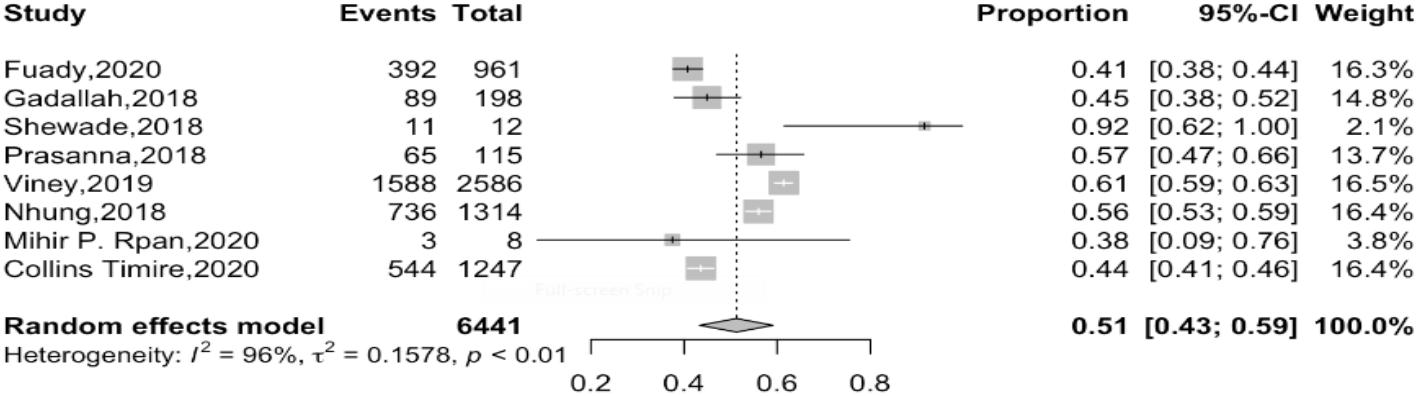
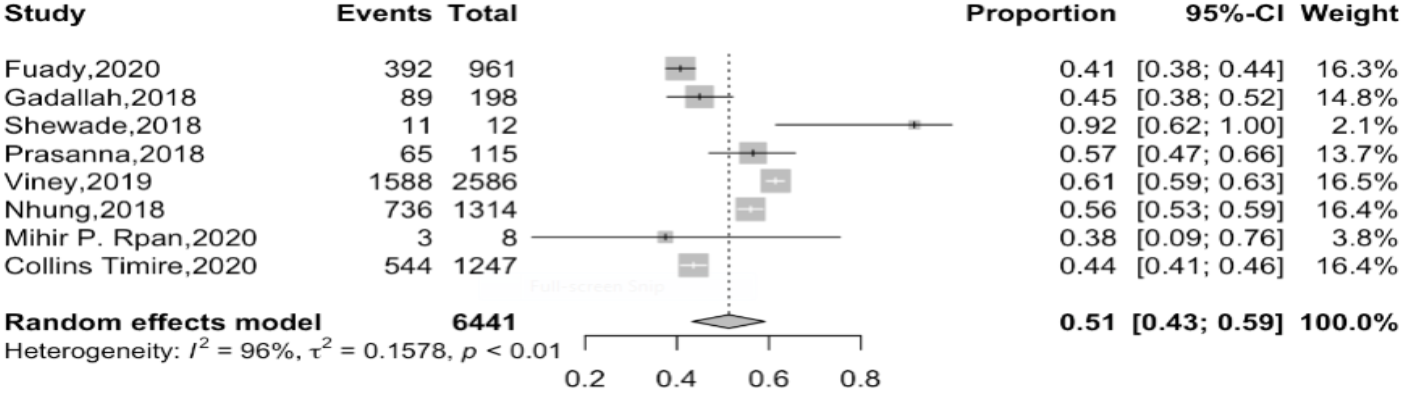
Pooled proportion of direct to total cost at catastrophic cost of 20%.

### 2.2. Catastrophic Health Expenditure at 10% & Capacity to Pay at 40%

In this study, we have found that there are six studies that also calculated the CHE 10% and the CTP 40%, in addition to their results regarding the CTC 20%.

#### 2.2.1. Pooled proportion of CHE at 10%

The pooled proportion of the CHE at 10% were studied also among the studies which they calculated CTC 20%. Three studies [2, 27, 32] were included with pooled proportion of 45%, 95%CI [35-56%], I^2^ = 93%. The result after leave one out sensitivity analysis, Fuady, 2018[<u>40</u>], has excluded and the heterogeneity has decreased to reach I^2^= 28%, while the pooled proportion has increased to 50%, 95%CI [47-54%]. **Figure (12)**

**Figure (12).**
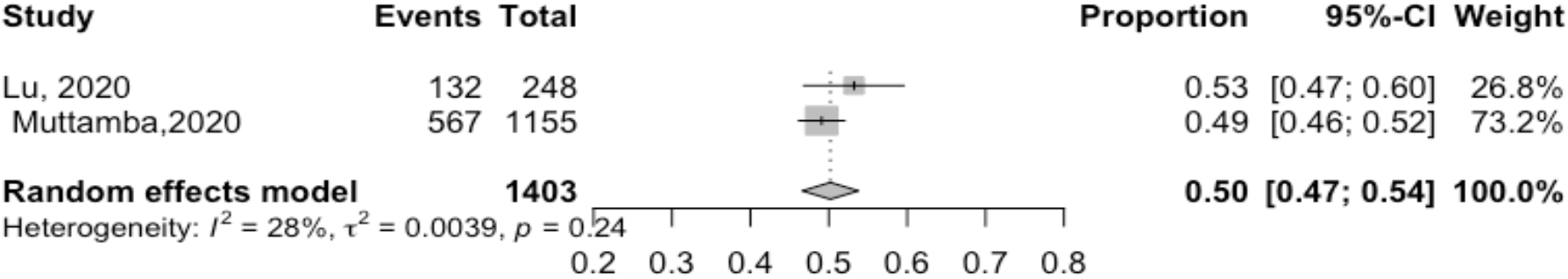
Pooled proportion of CHE at 10%.

**Figure (13).**
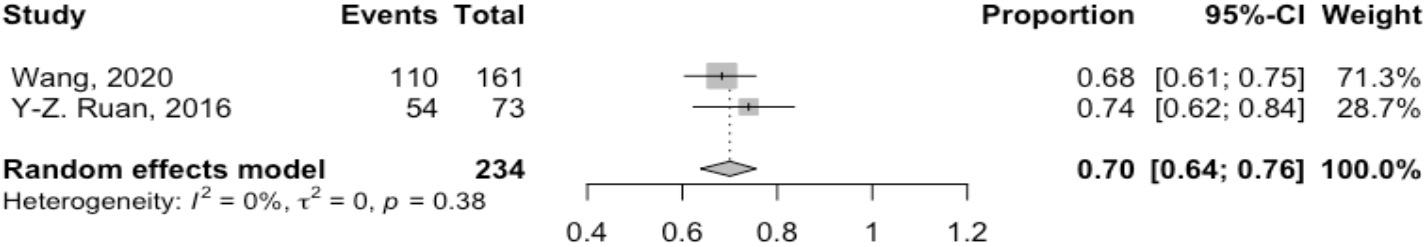
Pooled proportion of CTP at 40%.

#### 2.2.2. Pooled proportion of CTP at 40%

With 63% pooled proportion, 95%CI [40-80%], I^2^= 96%, three studies measured the CTP at 40% [12, 29, 30] and after the sensitivity test the heterogeneity was I^2^=0, while the pooled proportion increased to 70%, 95%CI [64-76%]. **Figure (15)**

## Discussion

Compared to the unknown data on the proportion of TB-patient affected household facing catastrophic cost in 2015, the GDGs goals set that 0% of household affected by TB have faced these costs by 2020 [51]. To the best of our knowledge, this is the first article that pooled of the proportion of TB patients or their households who suffered from catastrophic cost. In this meta-analysis 29 surveys conducted in 22 countries recruiting DS-TB, MDR-TB with or without HIV recruited through ACF, PCF. The quality score of the included studies ranged from 3-10. The proportion of patients facing catastrophic cost at a cut-off point 20% was 43%, (32%, 95%CI [29 – 35] among DS and 80% 95%CI [74-85%] among MDR). TB co-infected with HIV faced the highest catastrophic cost 81%, 95%CI [78 – 84]. Catastrophic cost was variables according to the strategy of case finding (ACF 12%95%CI [9-16%], versus PCF 42% 95%CI [35-50%]). The direct cost including medical and non-medical cost represented 51%, 95%CI [43-59%] of the total cost. Among drug sensitive and drug resistant TB, the proportion of direct cost to the total cost ranged from (33% to 65%)[15, 30, 33, 37, 41, 47] and (26%-93%)[15, 30, 32, 33, 37, 41, 47] respectively. ACF incurred lower catastrophic than PCF 29%, 95%C1 [20-41%] versus 37%, 95%C1 [34-40%]. The direct cost to the total cost among TB and HIV co-infected patients ranged from 30% [16]-59%[33]. The CHE was 50%, 95%CI [47-54%], and 70%, 95%CI [64-76%] at 10% of household yearly income and 40% of their capacity to pay respectively.

### Catastrophic cost

In fact, the cost incurred by some patients may be catastrophic and minimal for others. This is based on the household annual income. In the current study, we have included many studies that addressed the catastrophic cost among the TB at different thresholds, points (30%, 25%, 20%, 10% and 5%). Despite absence of robust evidence on the sensitivity of the cut-off point at 20% to reflect the catastrophic cost regardless patients are drug sensitive or resistant. Fuady et al, [12]settled 15% and 30% as more consistent cut-of points for treatment adherence and success respectively. In the current work, the proportion of TB-household patients facing catastrophic cost was 39%, which considered very high compared to the targeted GDGs in 2020 (0)%, more efforts and activities need to be directed to reduce this cost. It is worthy to note that diagnosis and treatment are provided for free in many of the included countries under the umbrella pooled of NTP, however, the treatment related expenditure is still very high. Yadav and his group, [52] illustrated that even with free services for tuberculosis care, 21.3% of the people in their study exposed to hardship financing, advising the need to take into consideration more innovated ways to increase the supported coverage of tuberculosis treatment in the country. The study also suggests the use of hardship financing as an index to measure the effectiveness of tuberculosis control program in the country. It is crucial to decrease the burden of catastrophic cost among the TB patients as it results in poorer treatment outcome. Patients suffer from catastrophic cost had 2-4 times higher odds of treatment failure than those who do not[12]. The latter is due to reduces access to the treating health facility, and treatment completion. Turning to the coping cost, a large proportion of household’s resort to different coping strategies to confront the increased out-of-pocket costs; and to compensate the consequences of income loss. Those coping strategies include selling a property or livestock, taking loans, pledging jewels, dropping their children out of school and cutting down their consumption to below basic needs [7]. Despite pooling of these studies’ outcome yielded substantial heterogeneity, the current study has found that almost 51% of heterogeneity, was mainly because of two predictors, the first was that some studies estimated CTC of DS and patients with MDR with or without HIV together. This factor played a major role in the heterogeneity, as it was clear that the CTC was dramatically higher among patients with HIV. The second predictor was the classification of country where the study was conducted[23]. Two-third of the new cases of TB reported in eight countries of the world, with India foremost the count, followed by Indonesia, China, the Philippines, Pakistan, Nigeria, Bangladesh and South Africa. Consequently, we divided studies into studies conducted in countries with high versus low incidence. In meta-regression, the country, where the study was conducted was a second major determinant of the different size effect.

The reported high incidence of CTC in many countries raised the need for social protection interventions. The most common social protection intervention is the cash transfer or cash assistance; it has already implemented in many countries across the world either conditionally or unconditionally [53]. In such a way, it is supposed that the household can get better access to treatment and food. Other social protection interventions include disability grants, food baskets (food assistance), food or travel vouchers and social insurance[7]. Many countries implemented reimbursement programs to help TB patients to cope with the disease cost. However, these programs prioritize poorer and MDR[54].The effect of this intervention is questionable. At a cutoff point of 20%, two studies have applied and calculated a catastrophic cost before and after reimbursement. Lue et al,2020[42] there reported a slight change on the proportion of CTC; before reimbursement, the CTC was (22%) and declined to 19% after the reimbursement. In contrary, Fuady,2019,[55] showed a higher change in the proportion of CTC after the reimbursement. The intervention program effectively decreased CTC from 44% to 13%. With regards to cash transfer, Wingfield et al, 2016 [56] reported that the proportion of TB household suffered from CTC was 30% and 42% among intervention and control respectively. These findings indicate that this social support is not enough to mitigate the impact of TB. Consequently, household of TB patients should receive sufficient financial support that covers the indirect cost (job lost), and direct cost (transportation, food, accommodation)[57].Of note, this social support should be proportionate to the income lost, this is due to the high variability of the pretreatment income. We speculate that development of newer treatment guidelines for TB of shorter duration would be beneficial. At the bottom, provision of free medication is not sufficient to prevent the catastrophic cost. TB patients should receive transport vouchers, reimbursement schemes and food assistance to reduce or compensate for such catastrophic costs. Furthermore, decentralization of patient supervision (including directly observed therapy), e.g. through community-based or workplace-based treatment [58], can reduce transport costs as well as income loss for patients[59].

As expected, the catastrophic cost among MDR was higher than DS, as DS patients receive treatment for shorter duration (6 months only), while MDR treatment extend to 24 months. Additional cost is incurred by MDR patients like the cost related to prolonged days of work absenteeism, need for daily injection, exposure to more side effects, and need for investigation [60].

### Direct cost to total cost

The total direct cost to the total cost was lower than the indirect cost among drug sensitive patients, HIV co-infected patients, while it was higher among drug resistant patients. This finding is essential to be considered when reimbursement strategies are implemented. Stakeholders should know which part of patient cost should be compensated. The direct cost dropped significantly if the strategy of active case finding was adopted instead of the passive case finding (29% to 37%) respectively.

### Determinant of catastrophic cost

Of note, it is essential to identify the factors that contribute to catastrophic cost. In this study, there are multiple main predictors of catastrophic cost. The main two components that affect the catastrophic cost are income loss as an impact of being diseased and food and nutritional supplements other than the patients’ regular diet habit addressing the catastrophic cost through increasing the direct non-medical costs [33–35]. Also travel and transportation affect the direct non-medical costs increasing the suffer of TB patients [30]. Age also considered to affect the prevalence of catastrophic cost whether the young age [27] or the old age [32].

### Catastrophic health expenditure

Out of the 29 studies, only six studies have been included with a clear measurement of the CHE at 10% of their income and 40% of their capacity to pay. It was clear that many studies ignored CHE, despite its importance to understand the impact of this cost on treatment outcome [42]. Two studies assessed the effect of reimbursements intervention on the CHE. Xiang et al, [61] reported a 8% reduction in CHE, however, this reduction was not statistically significant. Similarly, Zhou et al[62] reported that the effect of reimbursement on CHE was minimal, the achieved reduction in CHE was only 12%. In order to decrease the catastrophic expenditures National health financing systems must be designed and implemented, not to allow people to access services when they are needed only, but also to protect households from financial catastrophe, by reducing out-of-pocket spending. In the long run, prepayment mechanisms should be developed, for instance, social health insurance, tax-based financing of health care, or some mix of prepayment mechanisms such as efficient reimbursement or cash intervention. [63]

### Strength and limitation of the study

Our study has many strengths and limitations. Strengths include a comprehensive systematic approach to the existing literature, study selection, data extraction and quality assessment that have all been conducted according to current methodological standards. Furthermore, we included all studies without design, language, or geographical restriction. Moreover, we considered an ample list of outcomes and we compared these outcomes based on the definition, drug sensitivity and HIV infection. The limitation of this study was that different cut-off points were settled by different studies to estimate the proportion of the households facing catastrophic cost using different tools. A major challenge was that different studies estimated the catastrophic cost due to TB regardless drug sensitivity (DS, MDR), co-infection with HIV, case finding strategy (ACF, and PCF). Another point of limitation was that all studies included subjects with confirmed TB. Costs for those ill patients with undiagnosed TB may add a lot to the already estimated values. Furthermore, many of the included studies used the WHO cost survey tool, that include patients only treated in the NTP, omitting patients treated in private sectors who represent a considerable proportion of TB patients.

## Conclusion

About future global policy, our study provides evidence that despite the free TB treatment policy, there is a major proportion of TB patients are still facing catastrophic cost. The proportion of patient facing catastrophic cost is variable according to the type of TB; lowest among DS, higher in MDR, and highest if there is concomitant infection with HIV. Patients exposed to ACF incurred lower cost than those exposed to PCF. The direct cost (medical &non-medical) related to TB is not the only major contributor to the catastrophic cost, indirect cost represents a major contributor that should not be ignored. To sum up, this study paves the way to effective cost mitigation in the context of the End TB Strategy. As it addressed the proportion of TB patients and their households who are suffering from catastrophic cost and its predictors. Obviously, effective management of these predictors will eventually contribute to better community, clinical, financial outcomes [64]. Now it is clear that, the global health system must do more efforts to achieve the zero catastrophic cost for TB by 2030.

## Data Availability

all data are available.

## Acknowledgments

Many efforts overseas co-operate to finalize this work in this comprehensive way. We would like to thank Dr. Samia laokri and Dr. Charlesbatt, for giving us the opportunity to access their full-texts papers, which helped to make this Meta-analysis comprehensive, and guide us to the most robust outcome. Assistant Professor | Department of Instructional Technology and Learning Sciences, Otah University USA. We would like also, to express our indebtedness and deepest gratitude to Dr. Ramy shaaban, Dr. Suzan, and Dr. Ahmed Mandil (EMRO-WHO) for their great help, which enriched our search.

## Author contribution

**RMG**: Grant holder, conceptualized and designed the study, database search, full text screening, data analysis, writing manuscript.

**Haider El Saeh (HE):** Revision of tittle & abstract, revision of the full text screening and data extraction, data analysis and writing manuscript.

**Shaimaa Abdulaziz (ShA):** Database search with full text screening, data extraction, writing manuscript and references manager.

**Amira Elzorkany (AM):** Full text screening, data extraction, data analysis and writing manuscript.

**Nardin Zarif (NZ):** Database search with full text screening, data extraction and writing manuscript.

**Esraa Abdellatif (EAH):** Full text screening, data extraction and writing manuscript.

**Ehab Elrwiny (EE):** Full text screening, data extraction and writing manuscript.

**Samar Abdel-Hafeez (SA):** Final decision of the title & abstract screening with the full text screening, and writing manuscript.

